# TIME TO FIRST PASSAGE OF MECONIUM AND ASSOCIATED FACTORS IN 800 IRISH-BORN TERM INFANTS: A REAL-TIME OBSERVATIONAL STUDY

**DOI:** 10.1101/2025.06.12.25329211

**Authors:** A Byrne, G Avalos, A Rowan, A Waheed, NK Abidin, R O’Neill, E Kelly, A Corcoran, L Saba, E Moylett

## Abstract

**Objectives:** To develop an updated reference range for time to first passage of meconium in healthy, term, Irish-born infants and to determine associated obstetric and neonatal factors.

**Design:** A real-time observational study, conducted on the postnatal ward, to record the precise time of infants’ first passage of meconium. Associated obstetric and neonatal factors were analysed. Maternal consent was obtained prior to delivery. Frequency, percentages and median [IQR] values were reported. Non-parametric tests Mann Whitney U, Kruskall Wallis and Spearman Correlation, and Chi Square test and Fisher’s exact test were performed.

**Setting:** Level four, tertiary referral obstetric department

**Patients:** Healthy, term newborns.

**Main outcome measures:** Time to first passage of meconium, and obstetric and neonatal associations.

**Results:** In 800 infant subjects the median [IQR] time to first passage of meconium is 6.00 [9.45] hours (range 0-82.8 hours); 98.1% passed meconium within the first 24 hours. There was a statistically significant negative weak correlation between time to first passage of meconium and gestation, and between time to first passage of meconium and birth weight. Mean rank [median] time to first passage of meconium was greater and statistically significant for elective versus emergency caesarean section.

**Conclusions:** Ninety-eight percent of Irish-born newborns pass meconium in the first 24 hours. Meconium passage is influenced by gestation at delivery, birth weight, and emergency versus elective caesarean section.

**Key Messages:** *What is already known on this topic:* Most healthy infants will pass meconium within 24 to 48 hours of delivery. Up-to-date data are limited and heterogenous.

*What this study adds:* Despite changes in obstetric practice, most term healthy infants will pass meconium by 24 hours. Time to first passage of meconium is influenced by gestation and birth weight, and for those delivered by emergency versus elective caesarean section.

*How this study might affect research, practice or policy:* In the setting of modern obstetric care, infants who have not passed meconium by 24 hours should be clinically evaluated. Gestation and birth weight may be taken into account when assessing the timing of meconium passage.

## Background

Neonatal passage of meconium is a normal physiological event and an important indicator of newborn gastrointestinal function. Delayed or absent meconium passage requires urgent investigation and is often an indicator of underlying pathology, such as Hirschsprung disease, meconium ileus^1^ or anal atresia^2^.

A landmark study of 500 term infants by Sherry and Kramer (1955) found that almost 95% of babies will pass meconium by 24 hours, and over 99% by 48 hours^3^. Consequently, the timing of when to initiate investigations is typically after 24 to 48 hours of failure to pass meconium in term infants^4 5^. In the decades since, further data on time of first passage of meconium have been generated across different healthcare systems, in often heterogenous populations of newborns, primarily utilising retrospective data. Among studies based in high-income countries, a retrospective chart review by Metaj et al. of 1000 pre-term and term infants ≥34 weeks’ gestation (USA, 2003) found that over 90% passed meconium within the first 24 hours^6^. In the Netherlands, Bekkali et al. (2008) found that more than 79% of 71 term infants passed meconium within 24 hours and 83% within 48 hours, in a study comparing time of meconium passage in term and pre-term neonates^7^. In a low or middle income setting, Nigerian studies conducted in 2009^8^ and 2019^9^ found that only 75% and 56% of neonates, respectively, passed meconium within the first 24 hours.

Obstetric and postnatal practices have evolved significantly over the 70 years since Sherry and Kramer’s publication. Globally, caesarean section rates increased by 19 percentage points between 1990 and 2018, with an average of 21% globally and 27% in developed regions from 2010-2018^10^. In Ireland, the rate of caesarean section was 13-20% in the 1990s^11^. In 2023, this figure approached 37%, 38.3% at our institution^12^. Furthermore, global breastfeeding rates have increased since the 1980s^13^, including in Ireland^14^. However, a national study conducted in 2022 has shown that while 86% of babies born in Ireland were breastfed for their first feed, only 63% were exclusively breastfed in the first 48 hours of life^15^, and Irish breastfeeding rates are significantly lower now than in the 1960s, prior to the widespread availability of formula^14^. Finally, global fertility rates have decreased from 5.29 births per woman in the 1960s to 2.74 in the 2010s^16^. Studies into how these factors may influence timing of first meconium passage have yielded varied results.

Our study aimed to develop an updated reference range for time to first passage of meconium among healthy, term neonates, delivered in a modern, highly-resourced healthcare system, and explored potential influencing obstetric and neonatal factors. We sought to provide such reference data to assist with neonatal care, and to guide initiation of investigations into delayed passage of meconium.

## Methods

### Recruitment

Infants were recruited from a level four, tertiary referral obstetric department with 2,562 deliveries in 2020^17^. We recruited 800 healthy, term newborns delivered in our maternity department between October 2019 and September 2021, and September 2023 and February 2024. Recruitment was significantly impacted upon by the Covid-19 pandemic.

Recruitment was halted from December 2019 to September 2020, and operated at a reduced capacity from September 2020 to June 2021. Final recruitment took place from September 2023 to February 2024 to attain full power.

Newborns delivered between 37 and 42 weeks’ gestation, who did not require admission to the neonatal intensive care unit (NICU) were included; pre-term infants or those requiring NICU care were excluded. Infants of mothers requiring intensive care in the immediate post-partum period were also excluded. The minimum sample size of 785 infants was calculated using 80% power, p<0.05 level of significance.

Informed consent was obtained from expectant mothers on the antenatal and labour wards. During the consent process mothers were asked to record the date and precise time of their baby’s first meconium passage following delivery. If meconium-stained liquor was present or the baby passed meconium at delivery, this was recorded as time zero.

Owing to the nature of obstetric care, some mothers were unable to be recruited prior to delivery and therefore were not included, and of those recruited, some were unable to comply with study requirements and thus were excluded from analysis.

Ethical approval was obtained from a Clinical Research Ethics Committee, and human research ethics were adhered to throughout the study.

### Data collection

Time and date of first passage of meconium was recorded by the mothers or midwife at the time of consent. Data on obstetric and neonatal factors were retrieved from the maternal chart by data collectors using a standardised data collection form. Maternal data included age, parity, underlying medical conditions, medications, smoking status, alcohol consumption and use of recreational substances. Perinatal data included gestation, mode of delivery, use of anaesthetic agents, and other analgesia. Postnatal data included birth weight, time to first oral feed, type of first oral feed and infant ethnicity.

## Statistical analysis

Descriptive statistics were reported as frequencies and percentages, mean (SD) and median [IQR]. Data distribution suggested that time to first passage of meconium was not normally distributed, and non-parametric analyses were performed. The Mann-Whitney U and Kruskal-Wallis H tests were conducted to examine difference in mean ranks between time of meconium passage in relation to neonatal and obstetric factors. The results were presented as mean ranks [median, IQR]. Spearman’s correlation was used to find the correlation between time of first passage of meconium, and birth weight and gestation. The analysis of variance was used to analyse the difference in means amongst groups for other normally distributed continuous variables. Chi Square test and Fisher’s exact test were used to analyse neonatal and obstetric factors in infants who passed meconium before versus after 24 hours. While time to meconium passage was available for all 800 infants, some cases had missing data for obstetric and neonatal variables. Only cases with available data for those variables were included, to allow for maximum use of available data. Imputation was not performed for missing data. Data were analysed using IBM SPSS Version 29. Statistical significance was reported as p value less than 0.05. G*Power Version 3.1.9.4 was used to calculate sample size.

## Results

In total, 800 infants were recruited. Fifty-two percent (n=415) of infants were female, and 47.7% (n=379) were male. The median [IQR] gestation was 39 [1] weeks (n=260, 32.7%), and mean (SD) birthweight was 3,574 (466)gr. Four hundred and thirty (53.8%) infants were delivered via vaginal delivery, 369 (46.2%) by caesarean section; 200 (54.5%) of the latter group by emergency caesarean section. Ninety-three percent of infants (n=737) were Caucasian. Mean (SD) maternal age was 34 years, range 18-46. Median [IQR] parity was 1 [1], range 0-9. Further maternal and neonatal data are presented in Table 1.

**Table 1.** Demographic, Perinatal and Neonatal Data.

| **Variable** |  | **Results n (%)** |
| --- | --- | --- |
| Infant Gender | Female | 415 (52.3) |
|  | Male | 379 (47.7) |
| Gestation | 37 weeks | 53 (6.7) |
|  | 38 weeks | 103 (13.0) |
|  | 39 weeks | 260 (32.7) |
|  | 40 weeks | 234 (29.5) |
|  | 41 weeks | 133 (16.8) |
|  | 42 weeks | 11 (1.4) |
| Mode of Delivery | Caesarean Section | 369 (46.2) |
|  | Vaginal | 430 (53.8) |
| Category of Caesarean Section | Elective | 167 (45.5) |
|  | Emergency | 200 (54.5) |
| Infant Ethnicity | Caucasian | 737 (93.2) |
|  | Asian | 28 (3.5) |
|  | Mixed ethnicity | 15 (1.9) |
|  | Black | 7 (0.9) |
|  | Hispanic | 4 (0.5) |
| Maternal Parity | Primiparous | 345 (43.3) |
|  | Multiparous | 452 (56.7) |
| Anaesthetic in labour | Yes | 614 (77.1) |
|  | No | 182 (22.9) |
| Epidural in Labour | Yes | 349 (43.8) |
|  | No | 447 (56.2) |
| Mode of Feeding | Formula | 224 (28.3) |
|  | Breastmilk | 553 (69.8) |
|  | Mixed | 15 (1.9) |
| Time to First PO Feed | Within 1 hour of delivery | 591 (83.2) |
|  | >1 hour post delivery | 119 (16.8) |

Most of our cohort (n=785, 98.1%) passed meconium within twenty-four hours, and 99.5% (n=796) within forty-eight hours. The remaining four infants passed meconium up to 82.8 hours. The median [IQR] time to passage of first meconium (TPFM) was 6.00 [9.45] hours (min 0, max 82.8). Distribution of TPFM are detailed in Table 2 and Figure 1.

**Table 2.** Distribution of Time to Meconium Passage.

|  | *n* (%) |  | **Cumulative *n* (%)** |
| --- | --- | --- | --- |
| **<6.0 hours** | 398 (49.8) | **<6 hours** | 401 (49.8) |
| **6.0 – 12.0 hours** | 249 (31.1) | **≤12 hours** | 647 (80.9) |
| **>12.0 – 24.0 hours** | 138 (17.3) | **≤24 hours** | 785 (98.1) |
| **>24.0 – 36.0 hours** | 9 (1.1) | **≤36 hours** | 794 (99.3) |
| **>36.0 – 48.0 hours** | 2 (0.3) | **≤48 hours** | 796 (99.5) |
| **>48.0 hours** | 4 (0.5) | **Total** | 800 (100) |

Among those who passed meconium after 24 hours (n=15), 14 (93.3%) were breast-fed. Six (40.0%) were delivered by caesarean section; four emergency. Ten (66.7%) had spinal anaesthetic or epidural in labour and five (33.3%) had no anaesthetic. None required resuscitation at delivery. Five (33.3%) were fed within one hour of birth, ten (66.7%) over one hour after birth. All infants were discharged home following passage of meconium with no pathology identified and no intervention required.

Difference in proportions showed no significant association between; 1) Feeding type (breast 97.5% vs. formula 99.6%) and TFPM ≤24h vs >24h, χ^2^(1, N=777) = 3.66, p = 0.08; 2) Mode of delivery (Vaginal delivery 97.9% vs Caesarean section 98.4%) and TFPM ≤24h vs >24h, χ^2^(1, N = 799) = 0.235, p = 0.628; 3) Anaesthetic in labour 98.4% vs no anaesthetic 97.3% and TFPM ≤24h vs >24h, χ^2^(1, N = 796) = 0.950, p = 0.352; 4) Timing of first feed (less than one hour post-delivery 98.3% vs. greater than one hour 95.8%) and TFPM ≤24h vs >24h, χ^2^(1, N = 710) = 3.02 p=0.151.

There was a significant negative weak correlation in TFPM across gestational age groups, 37 to 42 weeks (rho= -0.193, p<0.001); later gestation was associated with shorter TFPM. (Figure 2)

Significant negative weak correlation was found between TFPM and birth weight (rho = - 0.120, p<0.001); increased birth weight correlated with decreased TFPM in this cohort. (Figure 3)

The mean rank [median] TFPM was not statistically significant for caesarean section compared to vaginal delivery (413 [6.31] vs 389 [5.88], p=0.135), however mean rank [median] TFPM was longer and weakly significant for elective (198 [6.90 hours]) versus emergency caesarean section (171 [5.59 hours]), p=0.017.

A positive very weak not significant correlation was found between TFPM and maternal age (rho=0.057, p=0.107); TFPM is greater with increasing maternal age.

The mean rank [median] for TFPM was not statistically significant for breast-fed versus formula-fed infants (397 [6.2 hours] vs 370 [5.5 hours], p=0.141). No statistically significant difference in mean ranks was found between time of first feed (within 1 hour of delivery vs greater than one hour) (p=0.957); epidural vs no epidural (p=0.088); infant gender (p=0.164), resuscitation at delivery vs no resuscitation (p=0.073).

## Discussion

Obstetric and maternal practices, including mode of delivery, infant feeding and birth rates, have evolved in the period since timing of first passage of meconium in term infants was initially studied. Research in this area has experienced heterogeneity in healthcare setting and study design. Our prospective, observational study of 800 infants is the first in over 60 years to investigate meconium passage in healthy, term infants in a developed healthcare system, and the first of its kind in an Irish hospital. The main findings of our study were: 1) median time to first passage of meconium was 6.00 hours, with 98% of infants passing meconium within 24 hours of delivery; and 2) time to passage of meconium is longer in infants of earlier term gestation, infants with smaller birth weight, and those born by elective versus emergency caesarean section. These findings would support updates to guidance on the timing of clinical review and investigation of infants with delayed passage of meconium (greater than 24 hours).

Our study found that 98% of infants passed meconium within 24 hours, and 99.5% within 48 hours. Similarly, Sherry and Kramer’s 1955 prospective study of 500 term, newborn infants, in Baltimore, USA, reported that 94% of infants first pass meconium within 24 hours of delivery^3^. Additionally, a 1977 prospective study of 500 term and pre-term infants by DA Clarke in Cleveland, USA^4^ identified that 99% of infants passed meconium by 24 hours. More recently, a 2003 retrospective study of 1,000 infants of ≥34 weeks’ gestation, by Metaj et al in Buffalo, USA^6^ reported that 97% of infants pass meconium within 24 hours. Despite significant changes in obstetric care over the past five decades^10-16^, our data support a similar time to first passage of meconium in healthy term infants born in highly resourced healthcare settings. Notably, data from Nigeria contrasts significantly, where between 56.6% and 75.3% of infants were reported to have passed meconium within the first 24 hours^8 9^. Thus, in developed healthcare settings, it is expected that most term infants will have passed meconium by 24 hours, and that failure to pass meconium by 24 hours should trigger clinical review and consideration of investigations. However, in low and middle income healthcare settings, further research may be of utility in defining an appropriate reference range for this cohort.

Numerous studies on both term and pre-term infants, conducted in African, American and Asian healthcare settings have shown that gestation is negatively correlated with time to meconium passage^9 6 18-21^. Among preterm infants, it has been shown in prior studies that gastric emptying and electrical activity are impaired^21-25^; manometric studies have identified antro-duodenal coordination immaturity and fewer duodenal clusters, which may contribute to the delay in first passage of meconium in this cohort^21 22 26 27^. However, our study has found that even among healthy, term infants of 37-42 weeks’ gestation, TFPM is longer for those of earlier gestation (rho= -0.193, p<0.001). Thus, even among term infants, for those born of full term or post-dates gestation, a lower threshold for investigation of delayed passage may be considered.

There is lack of cohesion in available data when comparing birth weight with time to first meconium passage. Previous studies which included both term and pre-term infants have demonstrated a negative correlation between birthweight and time to first meconium passage^9 19 20^. In contrast, two studies conducted in Nigeria, whereby pre-term infants were excluded and with sample sizes of 100 and 280 infants, found no such correlation^8 28^. Additionally, Bekkali et al reported in a 2008 prospective study of 198 Dutch-born infants, those who were ‘large for gestational age’, had a longer time to first passage of meconium than those with a normal birthweight^21^. Our study found that a lower birthweight correlates weakly with greater time to meconium passage (rho = -0.120, p<0.001). Given the heterogeneity in this data, further research is required before birthweight is considered in the clinical evaluation of an infant with delayed TFPM.

Regarding mode of delivery, a retrospective study of term and pre-term infants by Ezomike et al. found that time to meconium passage is significantly longer in infants born by caesarean section than those born by spontaneous vaginal delivery^9^. While our study found no statistically significant difference in mode of delivery, those born by emergency caesarean section were found to pass meconium more quickly than for elective caesarean section. This may be due to associated lower cord pH in infants born by emergency caesarean section versus elective^29^. Previously, Tateishi et al, proposed that the release of motilin and other gastrointestinal hormones, as a result of neonatal stress, may contribute to increased gut motility and therefore shorter time to meconium passage in infants born vaginally^30^. As our study sought to define a reference range for TFPM in healthy, term infants only, future research would benefit from the inclusion of term infants who required admission to the neonatal unit, as the impact of neonatal stress upon TFPM may be greater evaluated by including this cohort, when compared to a well population.

Published data on feed type and time to meconium passage is also varied with little consensus. Hameed et al. found that breastfed infants passed meconium sooner than formula-fed, in a prospective study of 222 term and pre-term neonates^19^. Like our own study, several other studies reported no such correlation^9 6 21 31^. However, our research recorded only the first feed following delivery. Future research which more broadly evaluates feeding type beyond first feed, to include all feeds within the first 24 to 48 hours, may better define the impact of breastmilk and formula on TFPM.

It is of crucial importance that babies with delayed passage of meconium are investigated in a timely manner. We would propose that healthy, term infants in highly-resourced healthcare settings who have not passed meconium by twenty-four hours should be carefully examined, particularly in the case of full-term and post-dates infants. Future research into infants who pass meconium after 24 hours, and in those who require neonatal unit admission in the postnatal period, may provide additional insights into the influence of individual obstetric and neonatal factors. Future studies should ensure that term and pre-term infants are considered as separate cohorts and are adequately powered as such. Recognition of differences in healthcare resource setting should guide local practice.

## Study limitations

This study relied on mothers to accurately record the time to first meconium passage of their newborns, and for these data to be collected by trained data collectors before discharge. Due to the Covid-19 pandemic, access to the maternity department by data collectors, and staffing levels were reduced. As a result, some data were missed in patients discharged before data could be collected. This resulted in an over-representation of infants delivered by Caesarean section, as these mothers remained inpatient for longer.

## Conclusion

Ninety-eight percent of healthy, term infants in a highly-resourced, modern healthcare setting pass meconium by twenty-four hours of life, and over ninety-nine by forty eight hours. Time to passage of meconium is significantly affected by gestation, birthweight and emergency versus elective caesarean section. These updated data may allow neonatal centres to develop stricter guidelines around reviewing and investigating infants who have not passed meconium by twenty-four hours, and may guide future research among term infants, particularly those who required admission to the neonatal unit.

## Supporting information

Supplement 2 Consent Form

## Data Availability

All data produced in the present study are available upon reasonable request to the authors

