## Supplement 1 Tables for "TIME TO FIRST PASSAGE OF MECONIUM AND ASSOCIATED FACTORS IN 800 IRISH-BORN TERM INFANTS: A REAL-TIME OBSERVATIONAL STUDY"

### **Supplementary Material 2**

### **Figure 1 Time to Meconium Passage Histogram**

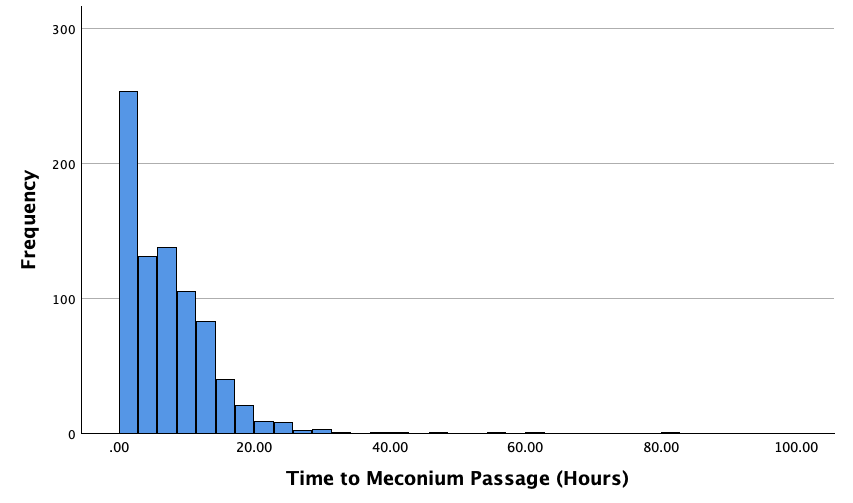

### **Figure 2 Correlation Time to Meconium Passage vs Gestation**

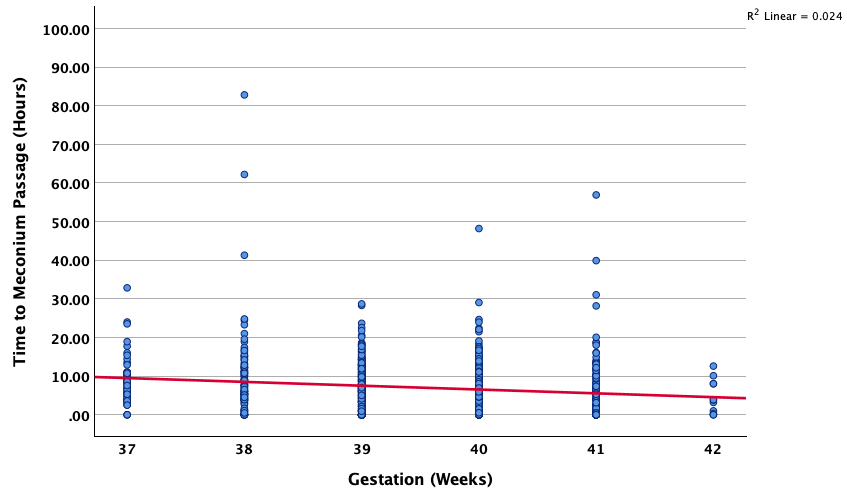

### **Figure 3 Time to Meconium Passage vs Birth Weight**

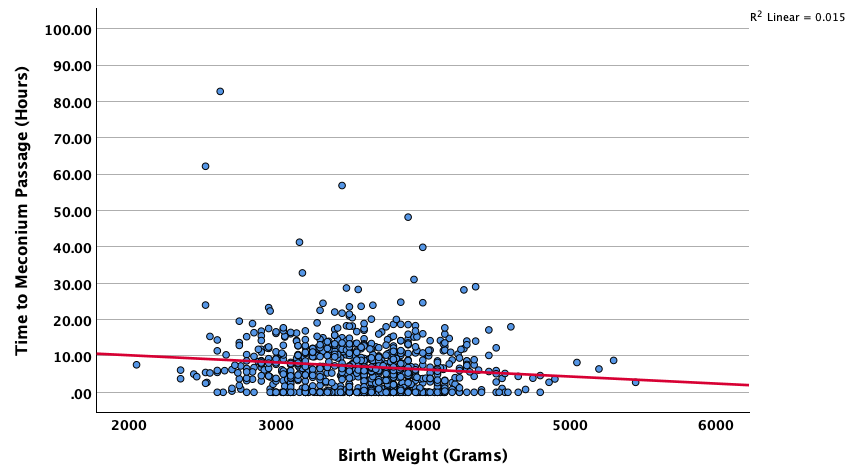
