## Supplement 2 Consent Form for "TIME TO FIRST PASSAGE OF MECONIUM AND ASSOCIATED FACTORS IN 800 IRISH-BORN TERM INFANTS: A REAL-TIME OBSERVATIONAL STUDY"

**Consent Form/Parent Information Sheet**

**September 2020**

**Title of Research Study:** Time to First Passage of Meconium in Irish Born Healthy Term Infants:

A Prospective Study

Academic Department of Paediatrics, NUI Galway

You are invited to **take part in a research study** under the guidance of the **Department of Paediatrics at the National University of Ireland Galway.** The purpose of the study is to **determine the exact time of a newborn baby’s first stool (meconium).** Most healthy babies will pass meconium in the first 48 hours of life.

We are undertaking this study because most of the medical literature on this topic is old (50 years ago) or the more current studies of a similar nature took place in the developing world.

By completing this study, we hope to determine a validated reference for average time to passage of meconium in healthy, term infants born in the developed world (Ireland).

In order to do this, we wish to record the **exact time following birth your baby passed meconium**. **Please record this time on the opposite side of this sheet**. We will use your medical record to gather information about your newborn including gestational age, birthweight, time of meconium passage, and the type and timing of first feed. Additionally we will use your medical record to obtain your age, how many babies you have given birth to, what type of delivery you had, if you were diagnosed with a medical condition in pregnancy, and any medications you were given (e.g. pain killers during labour).

Partaking in this study will not affect you or your baby’s care. The information will be anonymised, and we will only record your medical record number and your initials. After analysis is complete, all data will be destroyed.

**This study is entirely voluntary;** you are under no obligation to take part in this study. If you wish to take part, please sign below. If you have any questions, please ask the midwife looking after you or researcher as outlined below.

Thank you for your time and attention in reading this information sheet.

**Please give this completed form to the Midwife looking after you.**

THANK YOU

If you **AGREE** to partake in this study, please sign:

**Name: ______________________________________	Date: ________________________**

**Signature: ______________________________________	Your DOB: ____________________**

Principal Investigator:	Dr. Edina Moylett, Academic Department of Paediatrics, NUI Galway
